# Ophthalmological examination after hemispherotomy

**DOI:** 10.64898/2026.09.03.26362146

**Authors:** Thomas Schwartz, Vincent Toanen, Christine Bulteau, Dan Milea, Sarah Ferrand-Sorbet, François Audren

## Abstract

**Introduction:** Functional hemispheric disconnection (hemispherotomy) is an established surgical treatment for drug-resistant epilepsy. We aimed to describe visual function, including refractive errors, amblyopia, visual field defects, and oculomotor abnormalities, in a French cohort of patients who underwent hemispherotomy.

**Methods:** We retrospectively reviewed the ophthalmological records of all patients who underwent hemispherotomy at the Hôpital Fondation Adolphe de Rothschild between November 1987 and September 2024.

**Results:** Among the 368 patients who underwent hemispherotomy, 122 had a comprehensive ophthalmological assessment. Refractive errors were identified in 78 patients (66.7%). Amblyopia was diagnosed in 27 of the 89 patients in whom it could be assessed (30.3%). Of the 38 patients with reliable visual field testing, 37 (97.4%) had a homonymous hemianopia contralateral to the operated hemisphere. Oculomotor abnormalities were observed in 83 patients (68.0%), while 52 (42.6%) exhibited an abnormal head posture, suggesting compensatory mechanisms for visual field loss.

**Conclusions:** Ophthalmological abnormalities are highly prevalent following hemispherotomy. Systematic ophthalmological screening and appropriate management of treatable conditions, particularly refractive errors and amblyopia, appear warranted in this population. Patients also seem to develop compensatory oculomotor strategies and abnormal head postures in response to contralateral homonymous hemianopia.

## Introduction

Several aetiologies may lead to hemispheric epilepsy, including congenital neuronal migration disorders (such as focal cortical dysplasia and hemimegalencephaly) and destructive unilateral cerebral lesions (including Sturge–Weber syndrome, perinatal stroke, and Rasmussen encephalitis). Drug-resistant hemispheric epilepsy may require surgical treatment. Initially performed as anatomical hemispherectomy following its introduction in 1938, hemispheric surgery evolved during the 1990s with the development of hemispherotomy, a less invasive technique associated with fewer postoperative complications.(1)

Hemispherotomy consists of the functional disconnection of the affected cerebral hemisphere from the healthy contralateral hemisphere while preserving its vascular supply. The main indications for hemispherotomy are drug-resistant hemispheric epilepsies associated with hemiparesis or hemiplegia. Although patients usually experience persistent motor impairment and cognitive deficits following surgery, seizure freedom or a marked reduction in seizure frequency is associated with improved quality of life for both patients and their families, as well as better long-term cognitive outcomes.(2)

The literature on postoperative visual function following hemispherotomy remains limited. Koenraads et al. investigated visual outcomes after hemispherectomy. They reported postoperative homonymous hemianopia, which had already been present preoperatively in most patients, as well as clinical features suggestive of visual compensatory mechanisms, including exotropia and abnormal head posture.(3)

The aim of the present study was to describe visual function, including refractive errors, the prevalence of amblyopia, and oculomotor disorders, in a French cohort of patients who underwent hemispherotomy. Fundus examination and optical coherence tomography (OCT) findings were not investigated in the present study and are reported separately.

## Patients and methods

We retrospectively reviewed the medical records of all patients who underwent hemispherotomy at the Hôpital Fondation Adolphe de Rothschild between November 1987 and September 2024.

Patients were identified from the neurosurgical department database, from which demographic and clinical data were collected, including sex, date of surgery, age at surgery, epilepsy aetiology, side of hemispherotomy, cognitive status, and antiepileptic treatment at the time of the ophthalmological assessment. Intellectual functioning was evaluated using the Wechsler Adult Intelligence Scale, Fourth Edition (WAIS-IV), or other age-appropriate standardised neuropsychological tests.(4,5)

Ophthalmological data were retrieved from the Softalmo® electronic medical record system, used by the ophthalmology department since 2011, and from paper medical records for examinations performed before 2011. Ophthalmological assessments were performed by orthoptists and ophthalmologists. When multiple ophthalmological examinations were available, data from the most recent examination were analysed.

Visual acuity was measured using the Monoyer chart, recorded in decimal notation, and converted to logarithm of the minimum angle of resolution (logMAR) values for analysis. Refraction was assessed under cycloplegia using atropine 0.3% or 0.5%, according to patient age, or cyclopentolate 0.5%; when cycloplegia could not be performed, the fogging method was used instead. Refractive errors were classified according to the American Academy of Ophthalmology *Pediatric Eye Evaluations Preferred Practice Pattern* for patients younger than 12 years(6) and according to the criteria described by Williams et al. for patients aged 12 years or older.(7) The definitions used are provided in Appendix 1. The use of optical correction was recorded.

Amblyopia was defined as an interocular difference of at least two lines in best-corrected visual acuity and was classified as strabismic, anisometropic, mixed, or organic.

Whenever feasible, visual fields were assessed using Goldmann kinetic perimetry or Humphrey automated perimetry, depending on the patient’s ability to cooperate.

The presence and direction of abnormal head posture were recorded. Oculomotor assessment included ocular motility, prism cover testing when feasible, or corneal light reflex assessment when prism cover testing could not be performed. Ocular alignment (orthotropia, esotropia, or exotropia) and fixation preference were documented. Binocular vision was assessed using the Lang I stereotest.

An age cut-off of 3 years at surgery was defined *a priori* to distinguish procedures performed during the period of greatest cerebral plasticity. Patients were therefore divided into Group A (hemispherotomy performed at or before 3 years of age) and Group B (hemispherotomy performed after 3 years of age). Although this threshold does not represent a strict neurobiological boundary, it has been widely adopted in the hemispherotomy literature as a pragmatic clinical landmark, reflecting evidence that the capacity for functional reorganisation, particularly of the language systems, is greatest during early childhood.(8)

### Statistical analysis

All statistical analyses were performed using R® (version 4.5.2), and tables were prepared using Microsoft Excel®.

Univariable analyses were performed using non-parametric methods, namely Fisher’s exact test or the Cochran–Armitage trend test, as appropriate. Both unadjusted *p*-values and *p*-values adjusted using the Benjamini–Hochberg procedure were reported to control the false discovery rate (FDR) at 5%.

For the multivariable analyses, residual missing data in the explanatory variables were handled using multiple imputation by chained equations (MICE). Multicategory categorical variables were imputed using polytomous logistic regression, whereas binary variables were imputed using logistic regression. Binary logistic regression models were then fitted to each imputed dataset, and parameter estimates were pooled according to Rubin’s rules. Results are reported as odds ratios (ORs) with 95% confidence intervals (95% CIs).

The research was approved by the Rothschild Foundation Hospital review board – IRB 00012801-under the study number CE_20250923_3_TSZ.

## Results

Among the 368 patients who underwent hemispherotomy between November 1987 and September 2024, 146 underwent an ophthalmological assessment. Of these, 122 had at least one postoperative ophthalmological examination and were included in the study. The cohort comprised 63 females (51.6%), and the median age at surgery was 3.1 years. Fifty-five patients (45.1%) underwent right hemispherotomy. The underlying aetiologies were cerebral malformations in 58 patients (47.5%), Rasmussen syndrome in 29 (23.8%), stroke in 17 (13.9%), and Sturge–Weber syndrome in 18 (14.8%) (Table 1). Fifty-two patients (42.6%) had a normal intelligence quotient (IQ), whereas 23 (18.9%) had mild, 14 (11.5%) moderate, and 26 (21.3%) severe intellectual disability.

**Table 1.** Demography of the cohort.

| Aetiology | N (%) | Laterality of surgery (Right, %) | Median ±SD age at surgery (min; max) | Median ±SD age of last ophthalmological consultation (min; max) | Median ±SD interval between surgery and last ophthalmological consultation (min; max) |
| --- | --- | --- | --- | --- | --- |
| Stroke | 17 (14%) | 6 (35) | 2.9 ±4.5 (1.9;20.5) | 12.5 ±8.6 (3.5;29.7) | 7.8 ±7.7 (0.4;23.3) |
| Rasmussen syndrom | 29 (24%) | 12 (41.4) | 3.3 ±4.5 (3.3;16.7) | 11.4 ±8.4 (5.9;47.9) | 6.8 ±7.5 (0.4;31.1) |
| Cerebral malformation | 58 (47%) | 32 (55) | 3.1 ±4.7 (0.2;16.5) | 11.4 ±8.5 (0.4;36.5) | 7.1 ±7.5 (0;35.4) |
| Sturge Weber Krabbe syndrome | 18 (15%) | 5(27.8) | 2.9 ±4.6 (0.3;7.7) | 12.8 ±8.6 (1.3;26.9) | 8.0 ±7.7 (1.0;22.6) |
| Total | 122 (100%) | 55 | 3.1 ±4.6 (0.2;20.5) | 12.3 ±8.6 (0.4;47.9) | 7.3 ±7.7 (0.3;35.4) |

SD, Standard deviation

The median age at the last ophthalmological examination was 12.3 years, with a median postoperative follow-up of 7.3 years. At the time of the last ophthalmological assessment, 23 patients (18.9%) were still receiving antiepileptic medication.

### Refractive errors

Seventy-eight of the 117 patients with available refractive data (66.7%) had a refractive error classified as abnormal (Table 2 and Table 3). Cycloplegic refraction was performed in 51 patients. Overall, 81 of the 122 patients (66.4%) wore optical correction.

**Table 2.**
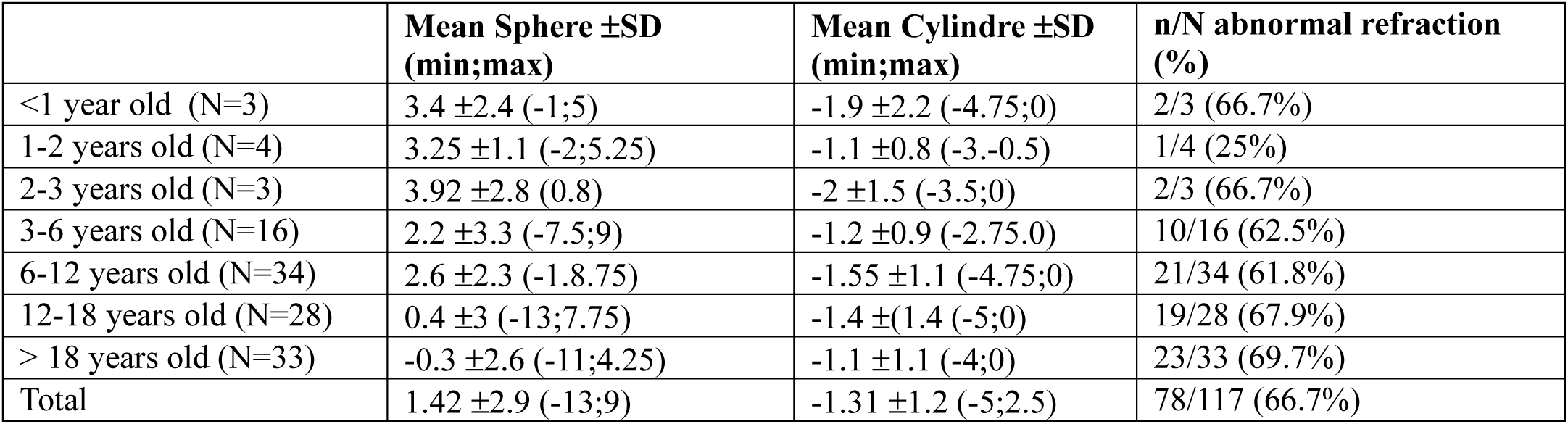
Mean spheric and cylindric refraction with standard deviation (SD), minimal and maximal values.

| | Mean Sphere ±SD (min;max) | Mean Cylindre ±SD (min;max) | n/N abnormal refraction (%) |
| --- | --- | --- | --- |
| <1 year old (N=3) | 3.4 ±2.4 (-1;5) | -1.9 ±2.2 (-4.75;0) | 2/3 (66.7%) |
| 1-2 years old (N=4) | 3.25 ±1.1 (-2;5.25) | -1.1 ±0.8 (-3.-0.5) | 1/4 (25%) |
| 2-3 years old (N=3) | 3.92 ±2.8 (0.8) | -2 ±1.5 (-3.5;0) | 2/3 (66.7%) |
| 3-6 years old (N=16) | 2.2 ±3.3 (-7.5;9) | -1.2 ±0.9 (-2.75;0) | 10/16 (62.5%) |
| 6-12 years old (N=34) | 2.6 ±2.3 (-1.8.75) | -1.55 ±1.1 (-4.75;0) | 21/34 (61.8%) |
| 12-18 years old (N=28) | 0.4 ±3 (-13;7.75) | -1.4 ±(1.4 (-5;0) | 19/28 (67.9%) |
| > 18 years old (N=33) | -0.3 ±2.6 (-11;4.25) | -1.1 ±1.1 (-4;0) | 23/33 (69.7%) |
| Total | 1.42 ±2.9 (-13;9) | -1.31 ±1.2 (-5;2.5) | 78/117 (66.7%) |

**Table 3.** Abnormal refraction: subtypes only for patients >12 years old.

| Refraction |  | n(%) |
| --- | --- | --- |
| Normal refraction |  |  |
| Hypermetropia N=42 |  |  |
|  | High hypermetropia | 7(16.7%) |
|  | Mild hypermetropia | 11 (21.2%) |
| Astigmatism |  | 58 (49.6%) |
| Myopia N=21 |  |  |
|  | High myopia* | 3(14.3%) |
|  | Mid myopia | 4 (19.0%) |
|  | Mild myopia | 12(57.1%) |

\*All 3 patients with high myopia had malformation, and 1 child with myopia had glaucoma with Surge-Webber.

Among patients with ametropia, 44 (56.4%) were in group A and 34 (58.6%) in group B (p = 0.23) (Table 4). The prevalence of ametropia according to underlying aetiology is presented in Appendix 2.

**Table 4.** Main outcome in group A (surgery before 3 years) and B (surgery after 3 years).

|  | Group A n/N (%) | Group B n/N (%) | p |
| --- | --- | --- | --- |
| Abnormal refraction | 44/59 (74.6%) | 38/58 (58.6%) | 0.006 |
| Amblyopia | 15/36 (41.7%) | 12/53 (22.6%) | 0.22 |
| Strabismus | 45/49 (91.8%) | 38/54 (70.4%) | 0.23 |
| Head turn | 29/35 (82.9%) | 24/31 (77.4%) | 0.86 |

### Visual acuity and amblyopia

Visual acuity could be reliably assessed in 89 patients. Of these, 27 (30.3%) were diagnosed with amblyopia. Fifteen (41.7%) patients in group A and 12 (22.6%) in group B were amblyopic (p = 0.22) (Table 4).

Among these patients, 16 (59.3%) had amblyopia on the side of hemispherotomy (p = 0.70) (Table 5).

**Table 5.**
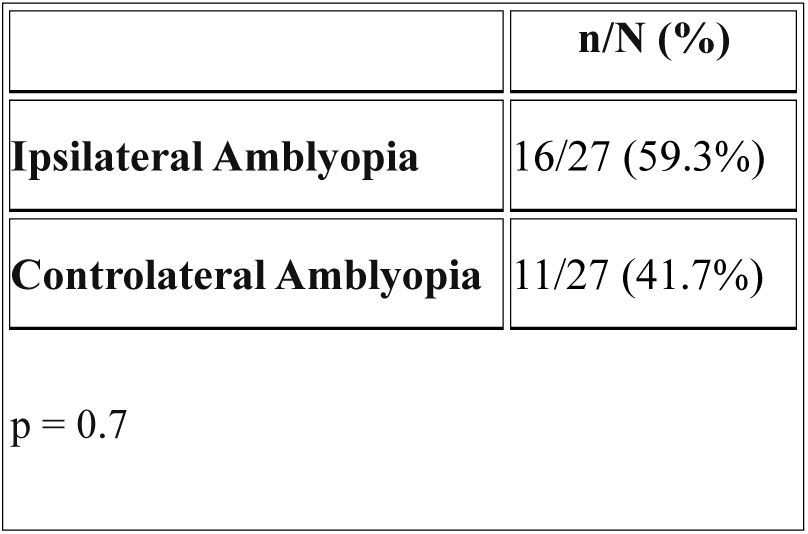
Amblyopia and side of surgery.

|  | n/N (%) |
| --- | --- |
| **Ipsilateral Amblyopia** | 16/27 (59.3%) |
| **Controlateral Amblyopia** | 11/27 (41.7%) |
| p = 0.7 |  |

Organic amblyopia predominated in patients with Sturge–Weber syndrome, accounting for six of the ten amblyopic patients in this subgroup (three had retinal detachment, one had multiply operated congenital glaucoma and had also undergone Descemet membrane endothelial keratoplasty (DMEK), one had undergone surgery for severe congenital glaucoma, and one had an ocular haemangioma). By contrast, among patients with cerebral malformations, strabismic amblyopia was most frequent, affecting six of the eleven amblyopic patients (the only case of organic amblyopia was secondary to retinal detachment). Refractive amblyopia was uncommon and was usually associated with either strabismus or an underlying organic visual pathway abnormality (Table 6).

**Table 6.** Types of amblyopia and aetiology.

| Aetiology | Amblyopia N (%) | VA max (logMar) | VA min (logMar) | Refractive amblyopia | Strabic amblyopia | Mixt amblyopia | Organic amblyopia |
| --- | --- | --- | --- | --- | --- | --- | --- |
| Malformation (N=58) | 11/58(19%) | 0 | LP- | 0 | 6 | 2 | 1 |
| Stroke (N=17) | 2/17 (12%) | 0 | 0.4 | 0 | 2 | 0 | 0 |
| Sturge-Weber (N=18) | 10/18 (55%) | 0 | LP- | 0 | 1 | 3 | 6 |
| Rasmussen (N=29) | 4/29 (14%) | 0 | 0.2 | 1 | 1 | 0 | 0 |
| Total (N=122) | 27/122 (22,1%) |  |  | 1 | 10 | 5 | 7 |

VA, Visual Acuity; LP-, Negative Light Perception

Ninety-four patients had never received occlusion therapy, whereas 18 had undergone patching treatment. Occlusion was applied to the right eye in 11 patients, to the left eye in one patient, and alternated between both eyes in six patients.

### Visual field

Thirty-eight patients were able to undergo visual field assessment. Of these, 25 underwent Goldmann perimetry and 13 Humphrey automated perimetry. All but one patient demonstrated a homonymous hemianopia (HH). The remaining patient, who had a cerebral malformation, exhibited bilateral tubular visual fields on Goldmann perimetry in the context of a right-sided Morning Glory optic disc anomaly and left peripapillary atrophy, suggestive of non-organic visual loss.

In all patients with homonymous hemianopia, the visual field defect was contralateral to the side of surgery, including 19 patients who underwent left hemispherotomy and 18 who underwent right hemispherotomy (p < 0.05) (Table 7).

**Table 7.** Visual field and aetiology.

| Etiology | Visual field n/N (%) |
| --- | --- |
| Stroke | 7/17 (41%) |
| Malformation | 13/58 (22%) |
| Sturge-Weber | 4/18 (22%) |
| Rasmussen | 19/29 (66%) |

### Strabismus

Strabismus was diagnosed in 83 (68.0%) of the 122 patients. Exotropia affected 62 patients, including 16 with intermittent exotropia. Nineteen patients presented with esotropia; among these, seven (36.8%) had significant hypermetropia. One patient had sensory strabismus (Sturge– Weber syndrome) and one had paralytic strabismus secondary to third cranial nerve palsy (Table 8).

**Table 8.** Type of strabismus and aetiology.

| **Strabismus** | **Stroke** | **Malformation** | **Sturge-Weber** | **Rasmussen** | **Total** |
| --- | --- | --- | --- | --- | --- |
| Exotropia | 5 | 24 | 4 | 13 | 46 |
| Intermittent exotropia | 4 | 5 | 4 | 3 | 16 |
| Esotropia | 5 | 13 | 0 | 1 | 19 |
| Paralytic strabismus | 0 | 1 | 0 | 0 | 1 |
| Sensorial strabismus | 0 | 0 | 1 | 0 | 1 |
| Total | 14 | 43 | 9 | 17 | 83 |

Thirty-eight (61.3%) patients demonstrated fixation preference of the eye ipsilateral to the hemispherotomy (p = 0.19) (Appendix 3).

Strabismus was observed in 45 of 49 patients (91.8%) in group A, compared with 38 of 54 patients (70.4%) in group B (p = 0.06) (Table 4).

Strabismus was present in 39 (81.3%) of 48 patients with a normal IQ and in 19 of 21 patients (90.5%) with severe intellectual disability. There was no significant association between cognitive status and the presence of strabismus (p = 0.64).

Only one patient underwent strabismus surgery in our institution, consisting of horizontal muscle surgery for symptomatic large-angle exotropia with diplopia. Two additional patients underwent exotropia surgery for cosmetic reasons.

### Abnormal head posture

Fifty-two patients exhibited abnormal head posture, including 48 with a constant head turn. In almost all cases, the head turn was directed contralaterally to the side of hemispherotomy (i.e. towards the side of the visual field defect), consistent with a compensatory mechanism. Among these patients, 42 (87.5%) had a contralateral head turn (p = 0.000003).

Only three patients with a right head turn had right-sided surgery (11.5%): one had tonic head and gaze deviation due to persistent seizures, one had esotropia with fixation in adduction, and one had previously undergone strabismus surgery and may not have had a true compensatory head posture (Appendix 4).

The prevalence of abnormal head posture according to underlying aetiology is shown in Appendix 5. Twenty-nine patients (82.7%) with head turn were in group A and 12 in group B (p = 0.22) (Table 4).

Twenty-two patients with abnormal head posture (44.9%) had associated intellectual disability, including seven with mild, two with moderate, and 13 with severe impairment. Abnormal head posture was present in 27 of 32 patients (84.4%) with normal IQ and in 13 of 15 patients (86.7%) with severe intellectual disability, with no significant association between cognitive status and abnormal head posture (p = 0.64).

### Multivariable analyses

Multivariable analyses did not identify any independent predictors of the outcome. Specifically, age at surgery, aetiology, refractive error, amblyopia, strabismus, and abnormal head posture were not independently associated with the outcome.

## Discussion

In this study, we report the prevalence of refractive errors, amblyopia, and oculomotor abnormalities in patients who underwent hemispherotomy. Ophthalmological abnormalities were common after surgery and included not only the compensatory oculomotor adaptations previously described, but also a high prevalence of refractive errors and amblyopia. In our cohort, 68% of patients had strabismus, almost half exhibited a compensatory head turn towards the side of the visual field defect, 66% had a refractive error, and 30% were diagnosed with amblyopia.

The prevalence of ametropia in our cohort appeared to be higher than that reported in the general population, although age-specific epidemiological data remain limited and comparisons between studies are difficult because of differences in definitions and study populations. Overall, two-thirds of our patients had at least one clinically significant refractive error. Children who underwent hemispherotomy before the age of 3 years appeared to be at greater risk, although this difference did not reach statistical significance. Likewise, patients in our cohort appeared more likely to require optical correction than individuals in the general population, particularly those with cerebral malformations, although this association was not statistically significant.

Twenty-seven patients were diagnosed with amblyopia. No significant association was found between the side of hemispherotomy and the side of amblyopia, although a trend towards ipsilateral amblyopia was observed among patients who underwent left hemispherotomy.

All but one patient who was able to undergo visual field testing demonstrated a contralateral homonymous hemianopia. This visual field defect is an expected consequence of hemispherotomy and was probably already present before surgery in many patients with these disorders causing hemispheric epilepsy, owing to early damage of the retrogeniculate visual pathways. Only a minority of patients were able to perform formal visual field testing, most of whom had Rasmussen encephalitis, an acquired condition in which visual fields are generally preserved before surgery and hemispherotomy is usually performed at an age when reliable perimetric assessment is feasible. In these patients, the visual field defect appeared to remain stable throughout follow-up.

Strabismus was present in 68% of patients, with a predominance of exotropia, which affected 51% of the overall cohort and accounted for 75% of all cases of strabismus. There was a non-significant trend towards ocular deviation contralateral to the operated hemisphere. These findings are consistent with those reported by Fujimoto et al.,(9) who also observed a predominance of exotropia.

The pathophysiology of postoperative strabismus remains incompletely understood. Chan et al.(10) provided evidence supporting a possible cortical contribution to strabismus. Using structural neuroimaging, they demonstrated that adults with strabismus exhibit reduced grey matter volume in the calcarine sulcus and occipital pole, as well as in the posterior intraparietal sulcus (IPS) and the right inferior parietal lobule (IPL), regions involved in visual motion processing, visuospatial integration, and depth perception. Conversely, increased grey matter volume was observed in cortical regions associated with oculomotor control. The authors proposed that compensatory ocular realignment in response to cortical visual deficits may promote adaptive structural plasticity within oculomotor networks. In contrast, Good et al.(11) suggested that postoperative exotropia is more likely to result from the underlying cerebral lesion than from adaptive cortical reorganisation.

Almost all patients with abnormal head posture exhibited a head turn contralateral to the operated hemisphere. The few patients with an ipsilateral head turn had plausible alternative explanations, including persistent seizures or fixation in adduction. The strong association observed for left-sided head turn, which remained significant after Benjamini–Hochberg correction, supports the hypothesis that compensatory head posture is directed contralaterally to the side of hemispherotomy, irrespective of the side of surgery. The absence of statistical significance for right-sided head turn is most likely explained by the limited sample size rather than by a true difference between right- and left-sided procedures.

Abnormal head posture also appears to represent a compensatory response to damage involving the frontal eye fields. Good et al. (11) proposed that head turning shifts the blind hemifield laterally, thereby optimising the patient’s functional visual field. In contrast, Paysse et al.(12) suggested that its primary purpose is to bring the intact visual field into a more central position. Koenraads et al.(3) further hypothesised that this dynamic adaptation facilitates active visual exploration of the environment. In their series, 38% of patients also exhibited contralateral exotropia. They additionally observed that compensatory adaptations were more frequently developed following right hemispheric surgery, possibly reflecting hemispheric specialisation, although the age at which the cerebral lesion occurred may also influence the extent of functional reorganisation.

Three patients in our cohort underwent surgery for large-angle exotropia. Given the likely compensatory role of both abnormal head posture and ocular deviation, strabismus surgery is generally discouraged in the literature. Nevertheless, none of the three patients, or their parents, reported any deterioration in visual function following surgery.

Age at hemispherotomy also appeared to influence the development of compensatory adaptations. Children who underwent surgery before the age of 3 years showed a trend towards a higher prevalence of postoperative strabismus, and a similar, although less pronounced, trend was observed for abnormal head posture. Although these associations did not reach statistical significance, they are consistent with the hypothesis that surgery during early childhood may favour the development of adaptive mechanisms through greater cerebral plasticity. In contrast, the severity of intellectual disability was not associated with the development of these compensatory adaptations.

### Strengths and limitations

The principal strength of this study is the size of the cohort. The Hôpital Fondation Adolphe de Rothschild is the national referral centre for hemispherotomy in France and manages almost all patients requiring this procedure nationwide, as well as a small number referred from neighbouring countries. To our knowledge, this is the largest ophthalmological series of patients undergoing hemispherotomy reported to date.

Several limitations should nevertheless be acknowledged. First, the retrospective design inevitably resulted in missing data. Ophthalmological assessment in this population is particularly challenging, both before and after surgery, because of neurological impairment, limited cooperation—mainly related to reduced attention span and fatigability—and the young age of many patients. To obtain reliable examinations, assessments often had to be spread over several visits, which was not always feasible. In addition, some patients lived far from the centre or abroad, making long-term ophthalmological follow-up difficult. Consequently, reliable assessment of ocular motility, visual acuity, or visual fields could not be obtained in all patients. Finally, this was a single-centre study. However, this limitation is partly mitigated by the fact that hemispherotomy is performed in only a limited number of centres in France, with specialised long-term ophthalmological follow-up concentrated in these centres. Therefore, our cohort is likely to be broadly representative of current national clinical practice.

### Conclusion

Ophthalmological abnormalities are highly prevalent following hemispherotomy. In particular, refractive errors, amblyopia, strabismus, and compensatory abnormal head posture are frequent findings. These results support the need for systematic ophthalmological assessment both before and after hemispherotomy, with long-term follow-up to enable early detection and management of treatable visual disorders, particularly refractive errors and amblyopia.

## Data Availability

All data generated during this study are available from the corresponding author upon reasonable request.

## Appendix

**Appendix 1.**
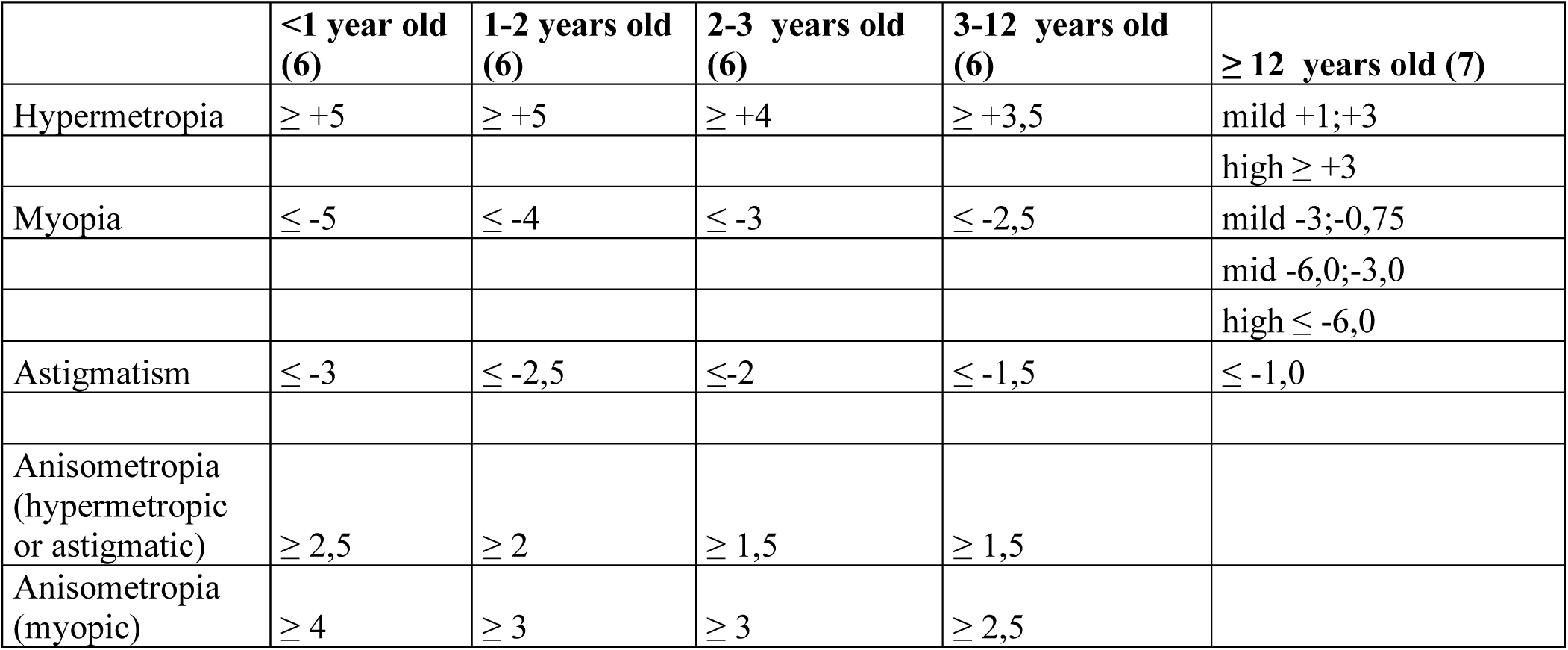
Summary of age-based refractive targets.

**Appendix 2.** Refractive error and aetiology.

| Aetiology | Refractive error n/N (%) |
| --- | --- |
| Stroke | 13/17 (76.5%) |
| Malformation | 40/55 (72.7%) |
| Sturge-Weber | 10/17 (58.8%) |
| Rasmussen | 15/28 (53.6%) |
| p = 0.52 |  |

**Appendix 3.** Strabismus and side of surgery.

|  | n/N (%) |
| --- | --- |
| **Ipsilateral fixation** | 38/62 (61.3) |
| **Controlateral fixation** | 24/62 (39.7) |
| P = 0.19 |  |

**Appendix 4.** Head turn and side of surgery.

|  | **n/N (%)** |
| --- | --- |
| **Ipsilateral head turn** | 6/48 (12.5%) |
| **Controlateral head turn** | 42/48 (87.5%) |
| p < 0.0001 |  |

**Appendix 5.** Abnormal head turn and aetiology.

| **Aetiology** | **Abnormal Head turn n/N (%)** |
| --- | --- |
| Stroke | 8/12 (66.7%) |
| Malformation | 27/33 (81.8%) |
| Sturge-Weber | 5/8 (62.5%) |
| Rasmussen | 12/14 (85.7%) |
| p = 0.72 |  |

